# Sewage, Salt, Silica and SARS-CoV-2 (4S): An economical kit-free method for direct capture of SARS-CoV-2 RNA from wastewater

**DOI:** 10.1101/2020.12.01.20242131

**Authors:** Oscar N. Whitney, Lauren C. Kennedy, Vinson Fan, Adrian Hinkle, Rose Kantor, Hannah Greenwald, Alexander Crits-Christoph, Basem Al-Shayeb, Mira Chaplin, Anna C. Maurer, Robert Tjian, Kara L. Nelson

## Abstract

Wastewater-based epidemiology is an emerging tool to monitor COVID-19 infection levels by measuring the concentration of severe acute respiratory syndrome coronavirus 2 (SARS-CoV-2) RNA in wastewater. There remains a need to improve wastewater RNA extraction methods’ sensitivity, speed, and reduce reliance on often expensive commercial reagents to make wastewater-based epidemiology more accessible. We present a kit-free wastewater RNA extraction method, titled “Sewage, Salt, Silica and SARS-CoV-2” (4S), that employs the abundant and affordable reagents sodium chloride (NaCl), ethanol and silica RNA capture matrices to recover 6-fold more SARS-CoV-2 RNA from wastewater than an existing ultrafiltration-based method. The 4S method concurrently recovered pepper mild mottle virus (PMMoV) and human 18S ribosomal subunit rRNA, both suitable as fecal concentration controls. The SARS-CoV-2 RNA concentrations measured in three sewersheds corresponded to the relative prevalence of COVID-19 infection determined via clinical testing. Lastly, controlled experiments indicate that the 4S method prevented RNA degradation during storage of wastewater samples, was compatible with heat pasteurization, and could be performed in approximately 3 hours. Overall, the 4S method is promising for effective, economical, and accessible wastewater-based epidemiology for SARS-CoV-2, providing another tool to fight the global pandemic.

**SYNOPSIS:** The 4S method for measuring SARS-CoV-2 in wastewater is promising for effective, economical, and accessible wastewater-based epidemiology.

**ABSTRACT ART:** 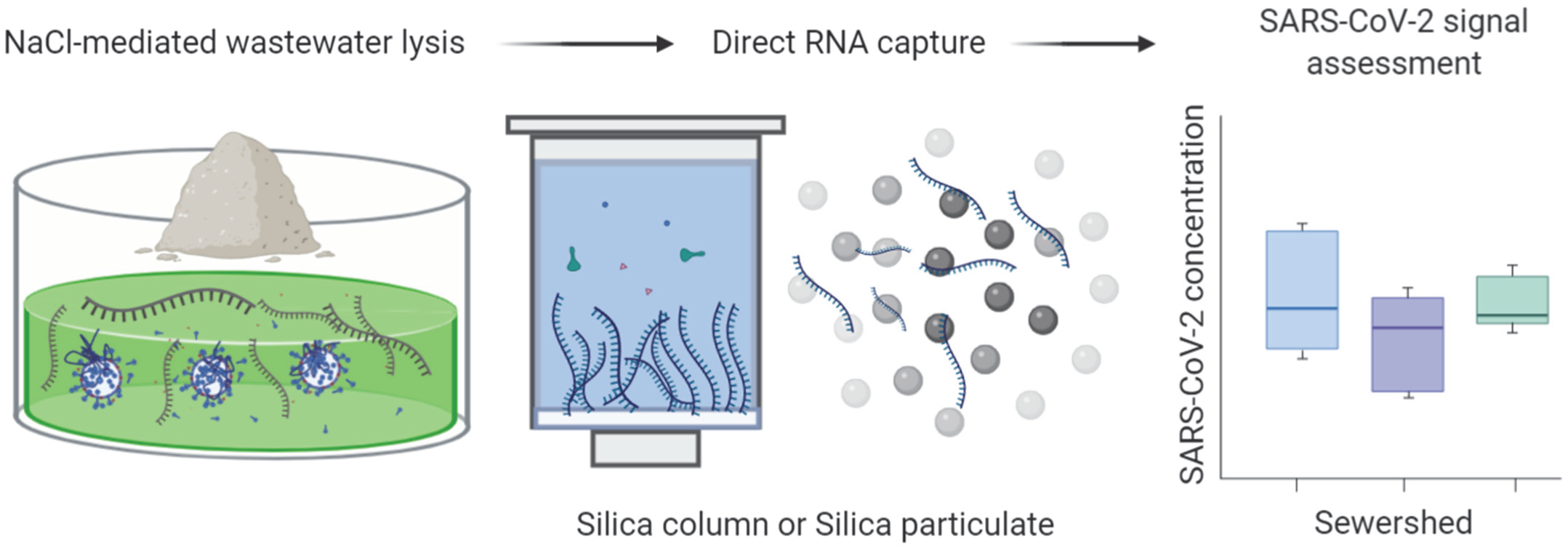

## INTRODUCTION

Wastewater-based epidemiology (WBE) enables the indirect assessment of viral infection prevalence in populations.^1–3^ The quantity of viral nucleic acids shed into wastewater by infected individuals, whether symptomatic or not, serves as a proxy for the relative prevalence of infection.^1^ WBE can provide population-level infection information for up to many thousands of individuals in a community to complement individual-level testing and aid public health decision making.^4^

WBE is now being applied to monitor and even predict population-level coronavirus disease 2019 (COVID-19) outbreaks.^1,5^ Local COVID-19 prevalence is difficult to assess due to insufficient individual testing capacity, rendering effective response more challenging.^6^ Wastewater can provide insights into COVID-19 prevalence, as COVID-19 patients shed SARS-CoV-2 RNA in their stool and thus into wastewater.^7,8^ Emerging studies report wastewater SARS-CoV-2 concentrations that correspond to reported clinical prevalence of COVID-19, with potential for early detection of COVID-19 outbreaks and identification of newly emerging SARS-CoV-2 variants.^9–12^ To extract and quantify the concentration of SARS-CoV-2 RNA shed into wastewater, researchers are using size- and charge-based concentration methods that concentrate intact SARS-CoV-2 virus prior to RNA extraction.^13–15^ These methods employ a primary concentration step via sieving by particle size, enmeshment of viral particles in precipitates that can be separated by mass, or adsorption via electrostatic interactions, prior to RNA extraction.^15^ These methods are relatively time-consuming and inaccessible as they are dependent on reliable supply of commercial reagents, a paucity of which has already hampered clinical SARS-CoV-2 testing efforts.^13–16^ Further, the use of primary concentration assumes the recovery of intact virus, and is therefore not geared towards co-capturing RNA from SARS-CoV-2 viruses that have already lysed or capture of non-viral RNAs suitable as fecal concentration controls.^14^ Lastly, current CDC safety guidelines recommend BSL-3 precautions when employing environmental sampling procedures that concentrate viruses presumed to be intact.^17^ To mitigate concerns of concentrating potentially infectious virus, heat-based wastewater sample pasteurization and subsequent extraction could allow for easier and safer wastewater processing after collection.

We aimed to develop an economical, kit-free method for the direct capture (extraction) of SARS-CoV-2 RNA from wastewater. The method employs lysis of biological particles via sodium chloride (NaCl), heat-based pasteurization, coarse filtration, ethanol precipitation, and RNA capture via silica-based columns (4S-column) or silicon dioxide slurry (4S-Milk-of-Silica). This approach allows recovery of wastewater RNA without mass, size, or charge bias and the co-capture of RNA from lysed SARS-CoV-2 virus alongside RNA from other biological particles in wastewater that are suitable as fecal concentration controls, such as pepper mild mottle virus (present in dietary peppers and shed in feces) and human 18S ribosomal RNA.^18^ The 4S method stabilizes the nucleic acids in wastewater via the addition of sodium chloride (NaCl) and ethylenediaminetetraacetic acid (EDTA), and is compatible with heat pasteurization, which makes wastewater samples safer to process. The 4S method’s omission of a primary concentration step and kit-free extraction enables lower reliance on commercial reagents and speeds up RNA purification to enable same-day measurement of SARS-CoV-2 wastewater abundance.

## MATERIALS AND METHODS

### Sample collection

For this study, we obtained composite 24-hour wastewater influent samples from East Bay Municipal Utility District’s wastewater treatment plant. These samples represent three discrete sampling areas: North and west Berkeley, El Cerrito, Kensington and Albany (sub-sewershed “N”), Oakland/Piedmont (sub-sewershed “S”), and Berkeley/Oakland Hills (sub-sewershed “A”) (interceptor coverage detailed in Fig. S2A). Samples were kept at 4°C on ice during transport and processed within 24 hours or kept at −80°C and processed within two weeks.

### Wastewater RNA extraction

Wastewater RNA extraction via the 4S-column and 4S-Milk-of-Silica methods is detailed in depth at https://www.protocols.io/view/v-4-direct-wastewater-rna-capture-and-purification-bpdfmi3n and dx.doi.org/10.17504/protocols.io.biwfkfbn.^28,29^. In brief, for 4S RNA extraction using a silica column, samples were lysed via the addition of Sodium Chloride (NaCl) to a final concentration of 4 M and EDTA to a final concentration of 1 mM and buffered via the addition of 10 mM pH 7.2 tris(hydroxymethyl)aminomethane. Samples were heat inactivated in a water bath (unless indicated otherwise) at 70°C for 45 minutes, filtered using a 5-μM DuraPore PVDF filter membrane (Millipore Sigma) and syringe filter. Ethanol was added to sample filtrate to a final concentration of 35%. Samples were passed through Zymo-IIIP silica columns (Zymo Research) using a vacuum manifold. For all experiments other than the wash buffer tests (Figure 4, Supplemental Figure S4), samples were washed with 25 mL of high NaCl (1.5M) and ethanol (20%) containing wash buffer #1 (4S-WB1), and 50 mL of low NaCl (100mM) and ethanol (80%) containing wash buffer #2 (4S-WB2). Washed RNA was eluted from silica columns using 200 μL of ZymoPURE elution buffer (Zymo Research) or pH 8 Tris-EDTA buffer pre-heated to 50°C.

For 4S-Milk-of-Silica extraction, samples were lysed, heat inactivated and filtered as in the 4S-column extraction. Next, a 1 g/mL silicon dioxide slurry in water was added to the filtered lysate and incubated at room temperature for 10 minutes. The lysate and silica slurry were centrifuged at 4000 x g for 5 minutes, pelleting wastewater RNA bound to silica particulate. The lysate supernatant was decanted, and the silica pellet was washed with 40 mL 4S-WB1 and 40 mL of 4S-WB2 via centrifugation and wash buffer decanting. The washed silica pellet was resuspended in 20 mL of pure water pre-heated to 37°C to elute bound RNA. Next, the silicon dioxide particulate was pelleted via centrifugation and the eluted RNA was separated and concentrated via isopropanol precipitation, as previously described.^30,29^ 4S-column and 4S-Milk-of-Silica reagent costs are listed in Supplementary Table 6.

For sample RNA concentration via ultrafiltration, Amicon 100-kDa ultrafilters (Millipore Sigma) were pretreated to block virus adsorption using 2 mL bovine serum albumen 1% (w/v) in 1x PBS and then washed with PBS. Wastewater samples were divided into 40 mL aliquots and solids were removed via slow centrifugation with a swinging bucket rotor at 4700 x g for 30 min. Supernatant was decanted and passed through a 0.2 µm flat membrane filter (Steriflip, EMD Millipore). Filtrate was loaded onto the ultrafilter in increments of up to 15 mL and ultrafilters were spun for 10 minutes at 4700 xg for each increment. Flow-through was discarded and samples were concentrated until they were reduced to a final volume of ∼250 µl. RNA was extracted from the ultrafiltration concentrate using an AllPrep DNA/RNA Mini kit (QIAGEN) following manufacturer instructions.

### RNA detection and quantification via RT-qPCR

This study employed four primer/probe sets: the SARS-CoV-2 N1 assay, Pepper mottle mild virus (PMMoV) coat protein gene assay, bovine coronavirus transmembrane protein gene assay and a newly developed human 18S ribosomal rRNA assay. (Supporting information, Table S3.) RT-qPCR reaction conditions are detailed in Table S1, assay thermocycling conditions are detailed in Table S2, and primer sequence information is in Table S3. RT-qPCR assay performance is detailed in Table S4 (Validation) and Table S5 (Limit of detection). RT-qPCR minimum information for publication of quantitative real-time PCR experiments (MIQE) documentation is detailed in Table S7. RT-qPCR analysis is detailed in the supporting information.

## RESULTS AND DISCUSSION

Many current methods of wastewater viral RNA extraction assume that most viral particles within wastewater are intact and that the concentration of these intact viruses prior to extraction is necessary to achieve sensitive detection of SARS-CoV-2 in wastewater. Given this assumption, these methods typically employ precipitation-, charge-, or size-based viral concentration and subsequent RNA extraction of unpasteurized wastewater to preserve viruses in an intact state.^14,15^ Despite concentration, some methods were shown to recover as little as 0-1% of SARS-CoV-1 from wastewater during the SARS-CoV-1 epidemic.^19^ We hypothesized that direct extraction could avoid loss of virus during the primary concentration step, and we therefore designed the 4S (Sewage, Salt, Silica and SARS-CoV-2) method to lyse viruses and microorganisms present in wastewater using sodium chloride and subsequently capture the free RNA using a silica RNA binding matrix.

To benchmark the performance of the 4S method, we analyzed a 24-hour composite wastewater sample treated with and without heat pasteurization and compared the recovery of endogenous SARS-CoV-2 to that of an ultrafiltration-based method. In addition, we compared the recovery of indigenous pepper mild mottle virus (PMMoV) RNA, which may be useful to control for variable fecal concentrations in wastewater, and a spiked-in bovine coronavirus vaccine (bCoV), used as an RNA extraction process control (Figure 1). We observed that the 4S-column method recovered 6-fold more SARS-CoV-2 RNA than ultrafiltration (Figure 1).

**Figure 1.**
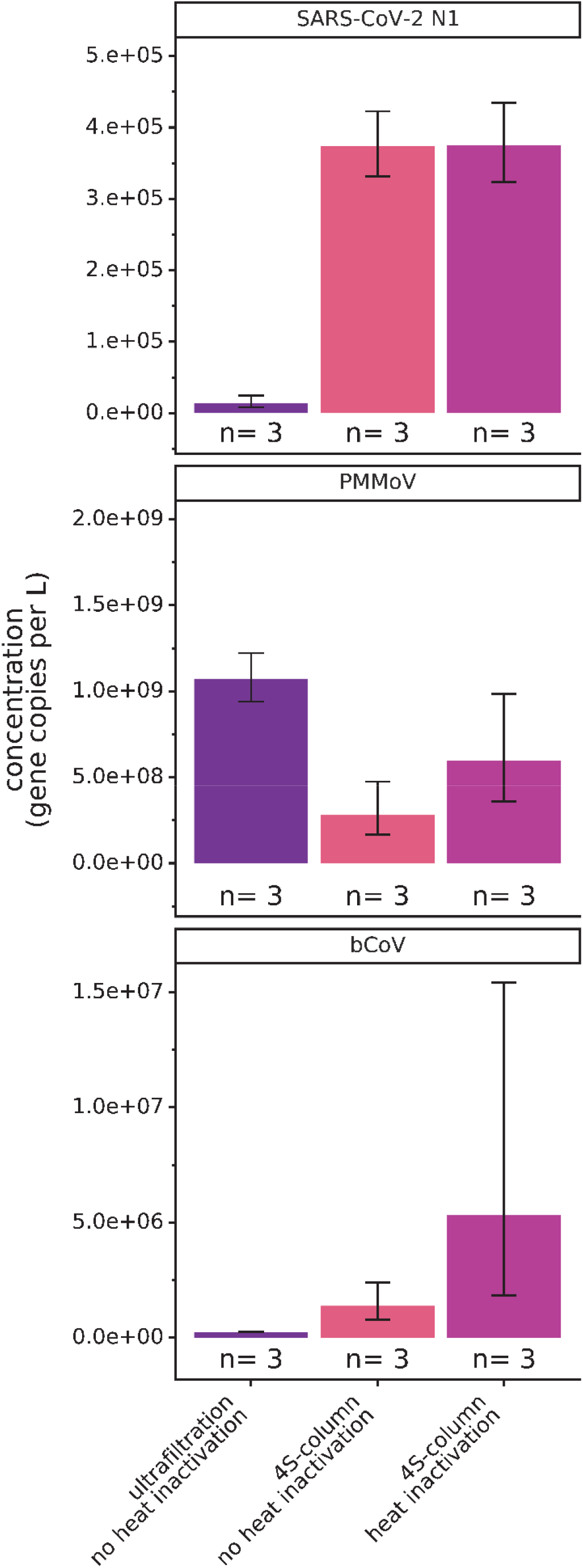
Comparison of SARS-CoV-2, PMMoV & bCoV spike-in assay signal in gene copies per liter between the 4S-column method with and without heat inactivation, and ultrafiltration. “n” represents the number of wastewater RNA extraction replicates per condition. Bars are plotted at the geometric mean of biological triplicates and error bars represent the variation associated with biological triplicates as quantified by the geometric standard deviation of the biological triplicates.

Surprisingly, SARS-CoV-2 recovery by the 4S method was not impacted by heat pasteurization, suggesting that further SARS-CoV-2 virus lysis did not occur. This result may imply that a large fraction of SARS-CoV-2 RNA was not bound to virus particles; this unbound RNA was captured by the 4S method but was not efficiently concentrated by ultrafiltration.

The 4S-column method without heat pasteurization also recovered 6-fold more bCoV than ultrafiltration, and 28-fold more bCoV with heat pasteurization. In this case, heat pasteurization may promote additional lysis of encapsidated bCoV, releasing its RNA for subsequent capture. Recovery of PMMoV by 4S was also higher with heat pasteurization (2-fold increase in recovery), but ultrafiltration was more effective in enriching PMMoV (1.6-fold higher than using the 4S-column method with heat pasteurization). Here, ultrafiltration may be effective in concentrating intact virus that is able to persist in wastewater, which is consistent with previous reports on PMMoV.^18,20^ Collectively, these results suggest that a significant fraction of SARS-CoV-2 RNA in the analyzed wastewater was not bound to viral particles but was present as free or ribonucleoprotein-bound RNA. This possibility is consistent with reports indicating reduced viability of SARS-CoV-2 and related coronaviruses spiked into wastewater.^13,21^

Given that the 4S method is designed to lyse and extract wastewater RNAs without requiring the enrichment of viral particles, we also investigated whether the 4S method could recover human RNAs present in wastewater. Using the 4S method, we were able to recover and detect human ribosomal subunit RNA (18S rRNA) in wastewater influent (Supplementary Figure S1A). 18S rRNA recovery was enhanced 2.5-fold by heat pasteurization, suggesting the lysis of human cells or 18S rRNA bound to ribonucleoprotein complexes present in wastewater (Supplementary Figure S1A). Therefore, the 4S method enabled the recovery and detection of human RNA, another potential indicator of wastewater fecal concentration, which could allow direct normalization of SARS-CoV-2 RNA quantity to human RNA content of wastewater. As heat pasteurization did not affect 4S recovery of SARS-CoV-2 and improved the recovery of PMMoV, bCoV and 18S rRNA, we recommend integrating this pathogen inactivation step to increase the safety of processing wastewater samples.

We sought to adapt the 4S strategy to employ silica powder for RNA capture rather than silica columns to circumvent reliance on commercially manufactured silica columns. In this approach, we added a slurry of silicon dioxide particles to lysed wastewater and used centrifugation to separate particle-bound RNA from the wastewater matrix, an approach we named “4S-Milk-of-Silica”. We observed that the 4S-Milk-of-Silica method recovered equivalent SARS-CoV-2 and PMMoV signal to the 4S method using a silica column (Supplementary Figure S1B). Thus, the 4S-Milk-of-Silica method presents an even more cost-effective (∼$8 per sample, vs. ∼$13 per sample, using the 4S-column extraction method, Supplementary Table 6) and accessible method to extract wastewater RNA without reliance on commercially manufactured silica columns and a vacuum manifold. However, the “Milk of Silica” version of the 4S protocol requires an isopropanol precipitation RNA concentration step, lengthening the protocol time. Therefore, we recommend using the 4S-column method to enable faster sample processing, while “4S-Milk-of-Silica” presents an alternate protocol for use in resource-limited settings.

WBE can provide an assessment of different areas’ relative COVID-19 infection prevalence, so we assessed whether the 4S-column extraction method could detect differential SARS-CoV-2 RNA levels in wastewaters derived from different subsections of a collection system. We surveyed three wastewater influent interceptors serving North and West Berkeley and El Cerrito (N), East Berkeley/Berkeley Hills (A) and Oakland (S) (Interceptor area coverage shown in Supplementary Figure S2A). These interceptors served areas exhibiting differential incidence of clinically confirmed COVID-19 cases, ranging from three (A interceptor) to 68 (S interceptor) reported cases per day within the week of our sampling (Figure S2A). To compare clinical COVID-19 case data and wastewater SARS-CoV-2 concentration, we normalized the case data by population, and we normalized the SARS-CoV-2 quantity by PMMoV abundance, to control for fecal concentration in the wastewater. Raw SARS-CoV2 and PMMoV abundance is available in Supplementary Figure 2B. As expected, the normalized wastewater SARS-CoV-2 signals trended with the per capita clinical cases per day in the three sub-sewersheds (Figure 2, B). The normalized SARS-CoV-2 RNA concentration was highest in wastewater representing the S interceptor area, where the highest daily per capita new cases also occurred. Normalized SARS-CoV-2 RNA concentrations in wastewaters representing the N interceptor area were only 2.3-fold lower than those of S interceptor wastewaters, despite 11.6-fold fewer per capita daily cases being reported in the A interceptor area during the week of our sample collection. One possible reason for this difference could be the presence of undiagnosed infections in the N interceptor service area, in which case wastewater SARS-CoV-2 RNA concentrations may provide a more accurate view of the relative COVID-19 infection prevalence in the week prior to sampling. Alternatively, the variability associated with wastewater measurements may be too large to detect differences of this magnitude.^9,13^ Ongoing research seeks to better quantify the measurement variability in wastewater samples over temporal and spatial scales. We emphasize that SARS-CoV-2 RNA levels were quantifiable in the A sub-sewershed despite only 18 cases being reported in an estimated population of 90,000 during the weeklong period of our sampling. This result implies that the 4S method is highly sensitive and can be used to monitor areas with low COVID-19 prevalence.

**Figure 2.**
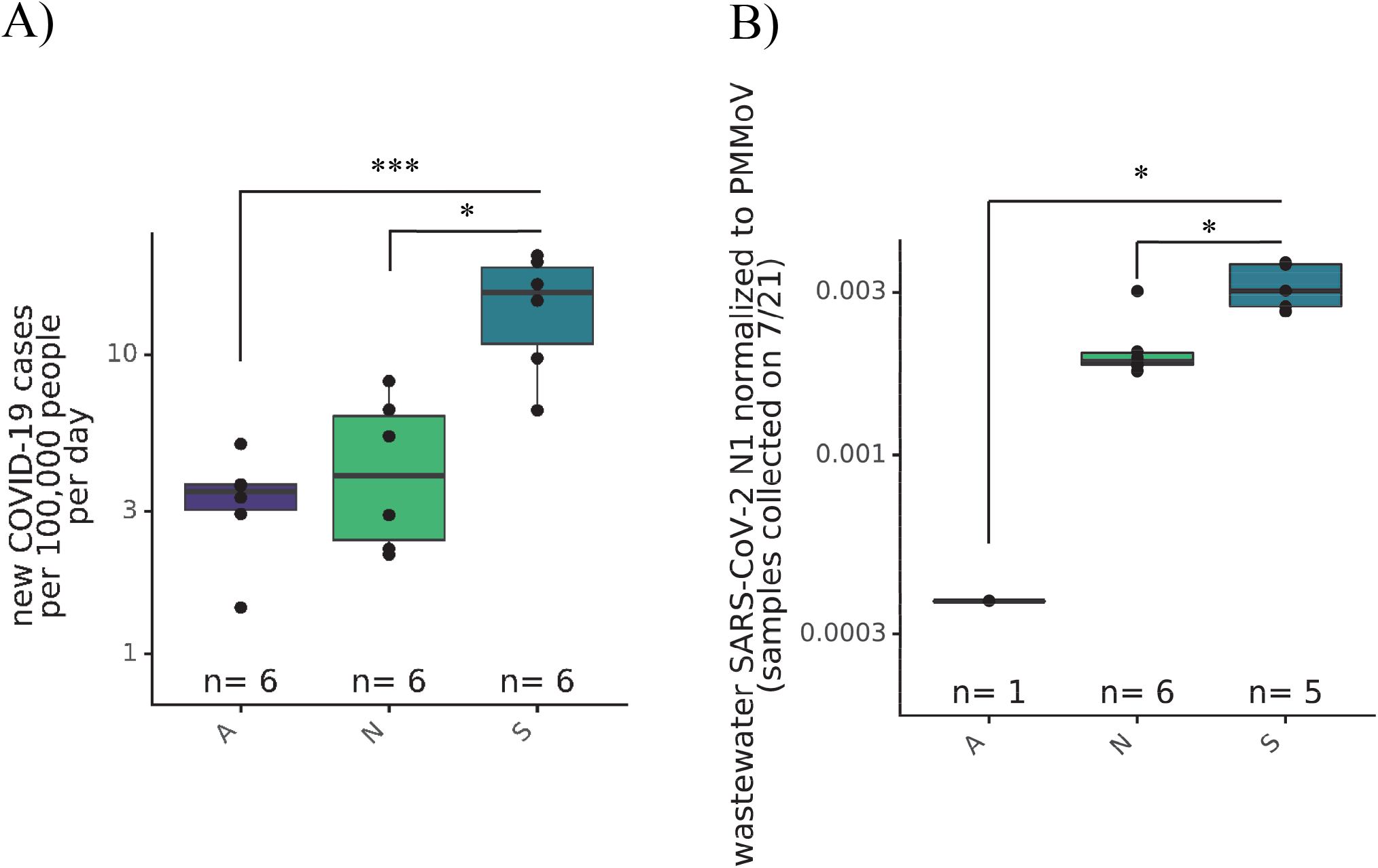
A) New COVID-19 cases per day per 100,000 population in three areas served by the distinct A, N and S wastewater interceptors over 6 days from 7/15 to 7/21. B) Comparison of SARS-CoV-2 N1 assay represented as SARS-CoV-2 gene copies per liter normalized to PMMoV gene copies per liter between interceptors serving the A, N and S East Bay areas. Kruskal-Wallis test followed by Dunn’s test was performed to determine significance, where *=p<0.05 and ***= p<0.001.

Wastewater contains many contaminants with the potential to degrade nucleic acids, and it has been previously observed that SARS-CoV-2 RNA in wastewater is degraded during storage.^22–24^ Viral detection relying on wastewater RNA extraction methods that concentrate intact viruses may be strongly affected by variable amounts of virus and viral RNA degradation in wastewater. Therefore, we sought to assess whether EDTA and sodium chloride, added to wastewater in the 4S method to promote lysis, could dually act to preserve RNA in wastewater. Upon receipt of each wastewater sample, we added NaCl to a final concentration of 4 M, added EDTA to a final concentration of 1 mM, and stored the samples either at 4°C for a month, or three days at room temperature (20°C). We observed that salt and EDTA addition prior to storage improved SARS-CoV-2 N1 assay signal after storage at both 4°C for one month (2.6-fold higher signal when stored with salt and EDTA) or at 20°C for three days (22-fold higher signal when stored with salt and EDTA) (Figure 3). Interestingly, the PMMoV assay signal remained similar throughout storage with or without salt, implying that PMMoV remains resistant to RNAses in the wastewater matrix. This observation corroborates previous reports indicating the persistence of PMMoV in wastewater.^18,20^ As with SARS-CoV-2 N1 signal, we observed that salt and EDTA addition preserved human 18S rRNA signal at 4°C for one month (126-fold higher) or at 20°C for three days (56-fold higher) (Figure 3). These results again support the conclusion that much of the SARS-CoV-2 in wastewater is not bound by intact capsid, rendering it more susceptible to degradation, unlike PMMoV which may remain encapsidated to protect it from degradation. Overall, the lysis salts added to wastewater as part of the normal 4S method workflow conveniently preserved wastewater RNAs and may mitigate degradation-mediated variation in SARS-CoV-2 and fecal concentration controls caused by RNA degradation during shipping and storage.

**Figure 3.**
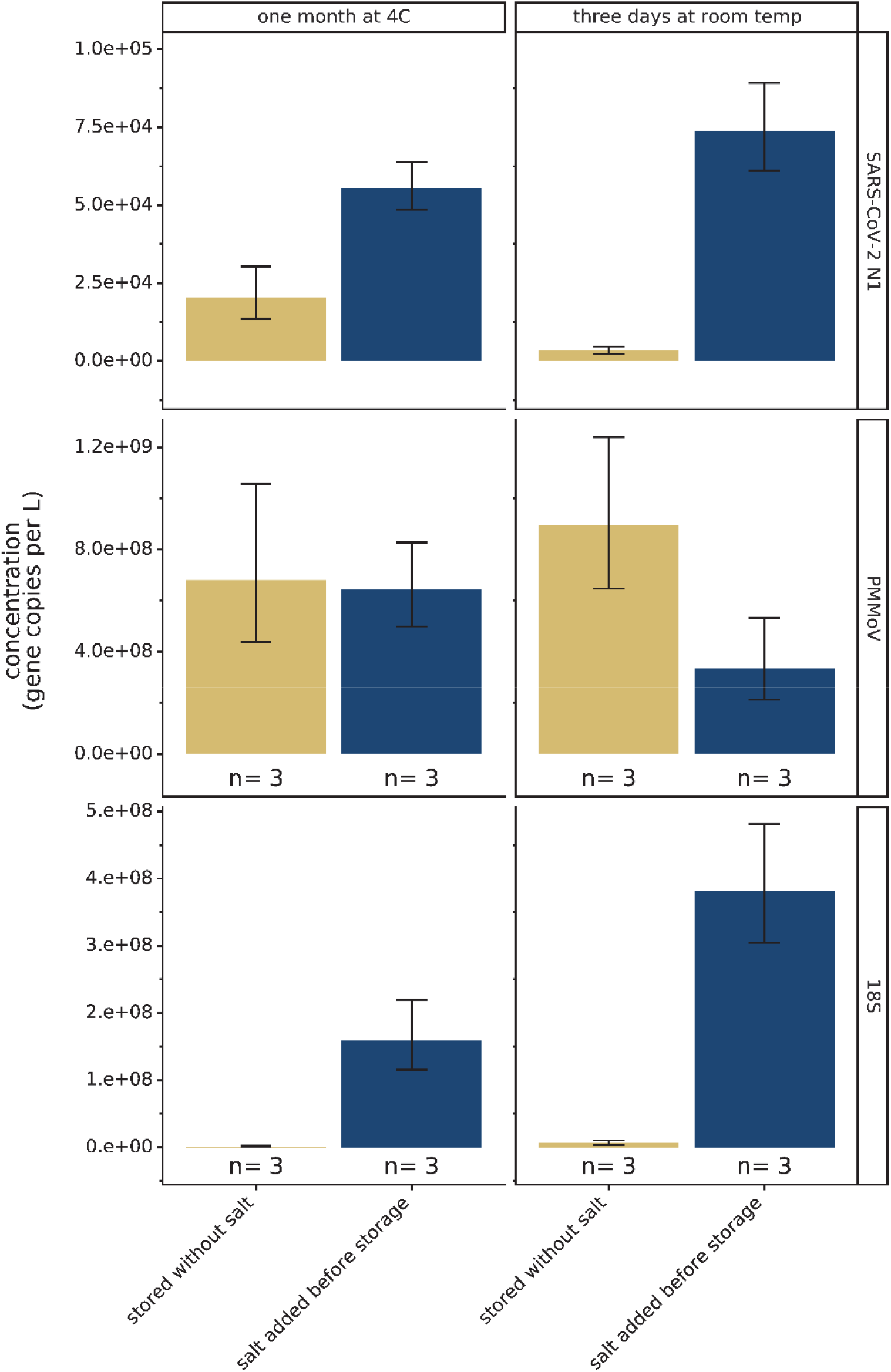
Effect of lysis salt addition prior to wastewater storage on SARS-CoV-2 N1, PMMoV and 18S rRNA assay signal. “n” represents the number of storage and extraction replicates per condition. Bars are plotted at the geometric mean of biological triplicates and error bars represent the variation associated with biological triplicates as quantified by the geometric standard deviation of the biological triplicates.

**Figure 4.**
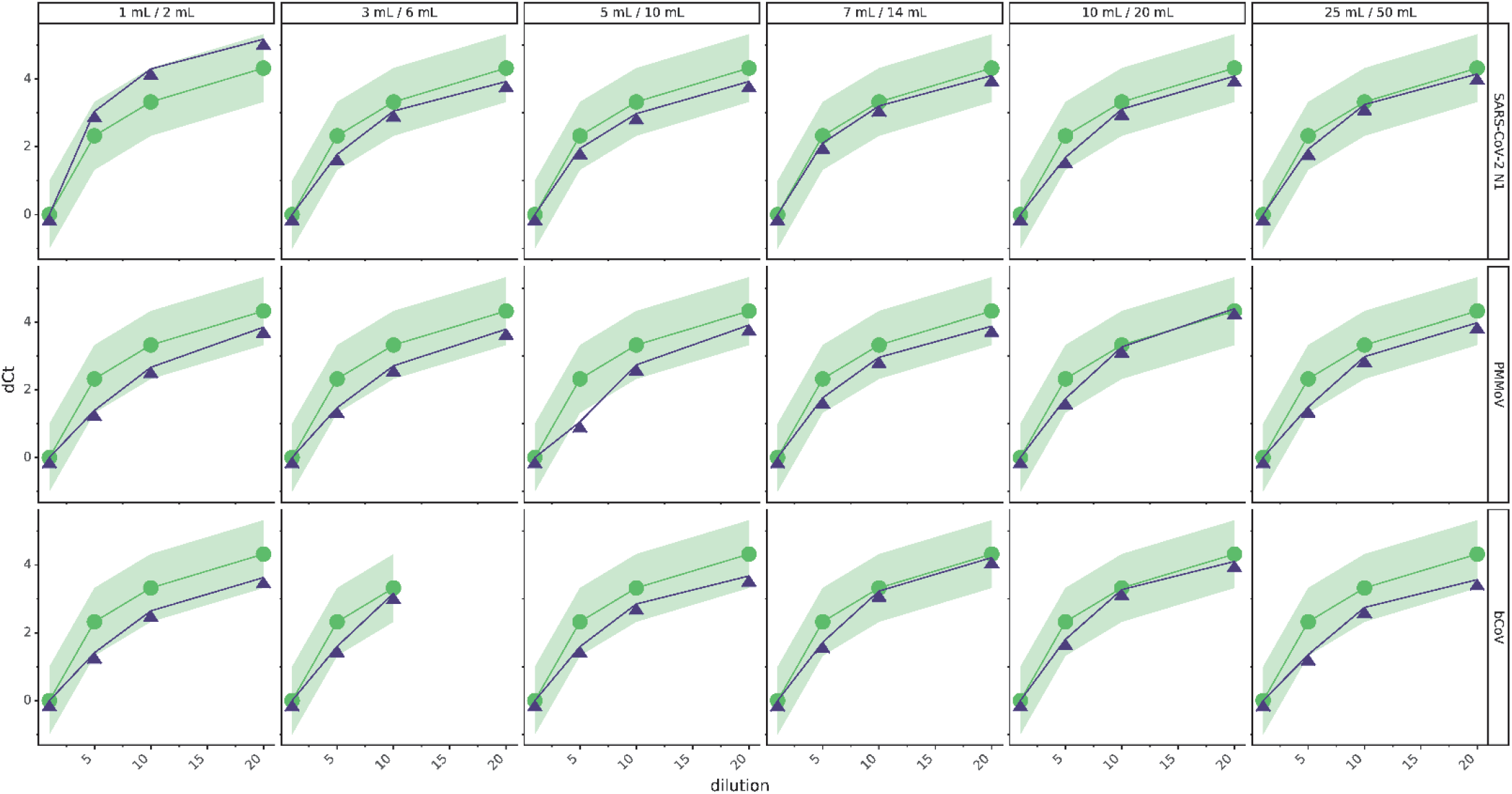
Assessment of RT-qPCR assay inhibition of the SARS-CoV-2 N1, PMMoV and bCoV assays via the “spike and dilute” method for different volumes of 4S-Wash buffer #1 and 4S-Wash buffer #2 (volumes reported at top of each panel). Sample dilutions shown are 1x, 5x, 10x, and 20x. Green line with circular points represents theoretically expected increase in Ct due to sample dilution, blue line with triangular points indicates actual increase in Ct with sample dilution. The green band indicates +/- 1 Ct tolerance range around the expected Ct values, due to variability. An increase in measured Ct that is lower than the expected increase was interpreted as inhibition. RNA sample dilution factor is indicated on x-axis.

Given the impact of RNA degradation on SARS-CoV-2 N1 assay signal, we investigated whether bulk RNA yield, representing intact wastewater RNA, could be employed as a normalization measure for SARS-CoV-2 detection. Surprisingly, bulk RNA yield per mL of wastewater input correlated poorly with SARS-CoV-2 and PMMoV detection (Supplementary Figure 3A). These results imply that most bulk wastewater RNA may be contributed by wastewater microorganisms unrelated to human fecal content or viral content, thus weakly correlating to SARS-CoV-2 N1 assay signal. We also observed that extracting nucleic acids from increasing volumes of wastewater (up to 400 mL) did not strongly increase total RNA yield per extraction past 100 mL of wastewater sample input, implying potential saturation of the RNA capture matrix (Supplementary Figure S3B). From these experiments, we conclude that the RT-qPCR detection of human fecal concentration indicators such as PMMoV and human 18S rRNA, the latter of which is preserved during storage similarly to SARS-CoV-2, are better estimators of wastewater fecal concentration than bulk RNA quantity measurements. Lastly, we observed that the 4S method enriched up to 8 µg of DNA per 100 mL of wastewater, suggesting that the 4S method could be employed for future wastewater surveillance of DNA-based pathogens and NA-sequencing based wastewater surveys (Supplementary Figure S3C).

Wastewater samples contain many contaminants that have previously been reported to inhibit RT-qPCR reactions.^25^ Therefore, we sought to assess whether the 4S method could generate purified RNA free of RT-qPCR contaminants by employing the “spike and dilute” method to assess PCR inhibition.^26^ Here, we spiked purified wastewater RNA with synthetic RNA standard and sequentially diluted the sample and observed whether SARS-CoV-2 N1, PMMoV, and bCoV detection followed corresponding sample dilutions, indicating an absence of inhibition. We assessed the impact of a range (1-50 mL) of wash buffer volumes during RNA extraction on PCR inhibition and SARS-CoV-2 N1, PMMoV, and bCoV assay signal to identify the optimal wash buffer volume for RNA purity and recovery. There was no evidence of inhibition for the SARS-CoV-2 N1 assay using the 4S procedure with any wash buffer volume, and slight inhibition of the PMMoV assay when using 5 mL of 4S-Wash buffer #1 (4S-WB1) and 10 mL of 4S-Wash buffer #2 (4S-WB2) (Figure 4). To limit ethanol waste generation, we therefore recommend using at least 7 mL of 4S-WB1 and 14 mL of 4S-WB2 to yield inhibitor-free RNA.

Next, we assessed potential assay signal loss due to excess washing of the silica columns. Here, we observed highest SARS-CoV-2, PMMoV and bCoV assay signal using 3 mL of 4S-WB1 and 6mL of Wash 4S-WB2, with minimal losses in signal up until 25 mL of 4S-WB1 and 50 mL of 4S-WB2 (Supplementary Figure S4). Using too little wash buffer may not sufficiently wash away lysis salts and contaminants from the silica matrix, reducing RNA recovery and increasing inhibition, whereas too much wash buffer may partially elute bound RNA, decreasing RNA yield. Therefore, we recommend using 7-10 mL of 4S-WB1 and 14-20 mL of 4S-WB2 to extract PCR inhibitor-free RNA while maximizing target RNA recovery.

The results presented here are representative of only three wastewater sources which may differ in composition from wastewater collected at other times and from other locations. Different wastewaters may contain different types and quantities of PCR inhibitors, so we recommend assessing PCR inhibition in all sample types, and if necessary, adjusting the wash buffer volumes to effectively remove inhibitors from the purified RNA. Different wastewater samples may also contain varying biological and chemical species influencing RNA stability, potentially impacting the RNA preservation documented here by the 4S method. Furthermore, the 4S method may be less effective in capturing the nucleic acids from wastewater viruses or other microorganisms resistant to the sodium chloride and heat-based lysis evaluated here.

Overall, we demonstrate that the 4S method enabled efficient extraction of SARS-CoV-2, PMMoV, bCoV, and human 18S rRNA. Combined with RT-qPCR, the 4S method allowed monitoring of relative COVID-19 infection prevalence with high sensitivity. These results are consistent with those of a recent inter-laboratory comparison of 36 different wastewater SARS-CoV-2 RNA detection methods. In this comparison, the concentration of SARS-CoV-2 measured with the 4S method, identified as “1S.2H”, was one of the highest reported (direct measurement, without correcting for recovery efficiency) and the recovery efficiency of a spiked-in OC43 virus efficiency control was the highest reported, among all methods.^27^ The 4S method also preserved RNA in wastewater, was compatible with heat pasteurization, and yielded purified RNA free of PCR inhibitors. Given the high efficiency, low cost, and same-day assessment of wastewater SARS-CoV-2 and fecal concentration controls, the 4S method presents an affordable and accessible method for implementing wastewater-based epidemiology for SARS-CoV-2. The method also appears promising for the application of WBE for other RNA- and DNA-based pathogens and facilitating research on the wastewater microbial community more broadly.

## Data Availability

All unprocessed data are available upon request.

## Acknowledgements

We thank all members of the Nelson laboratory and UC Berkeley COVID-WEB pop-up lab for their contributions, reading, insights and helpful discussions of this paper. We also thank all members of the Tjian and Darzacq lab for their support designing and carrying out molecular assays and experiments. We thank the East Bay Municipal Utility District for sample collection.

## References

1. Bivins, A. et al. Wastewater-Based Epidemiology: Global Collaborative to Maximize Contributions in the Fight Against COVID-19. Environ. Sci. Technol. 54, 7754–7757 (2020).

2. Zhou, N. A. et al. Feasibility of the Bag-Mediated Filtration System for Environmental Surveillance of Poliovirus in Kenya. Food Environ Virol 12, 35–47 (2020).

3. Kazama, S. et al. Environmental Surveillance of Norovirus Genogroups I and II for Sensitive Detection of Epidemic Variants. Appl. Environ. Microbiol. 83, (2017).

4. Kaliner, E. et al. The Israeli public health response to wild poliovirus importation. The Lancet Infectious Diseases 15, 1236–1242 (2015).

5. Hata, A. & Honda, R. Potential Sensitivity of Wastewater Monitoring for SARS-CoV-2: Comparison with Norovirus Cases. Environ. Sci. Technol. (2020) doi:10.1021/acs.est.0c02271.

6. Li, R. et al. Substantial undocumented infection facilitates the rapid dissemination of novel coronavirus (SARS-CoV-2). Science 368, 489–493 (2020).

7. Wu, Y. et al. Prolonged presence of SARS-CoV-2 viral RNA in faecal samples. The Lancet Gastroenterology & Hepatology 5, 434–435 (2020).

8. Holshue, M. L. et al. First Case of 2019 Novel Coronavirus in the United States. New England Journal of Medicine 382, 929–936 (2020).

9. Peccia, J. et al. Measurement of SARS-CoV-2 RNA in wastewater tracks community infection dynamics. Nature Biotechnology 38, 1164–1167 (2020).

10. Ahmed, W. et al. First confirmed detection of SARS-CoV-2 in untreated wastewater in Australia: A proof of concept for the wastewater surveillance of COVID-19 in the community. Science of The Total Environment 38764 (2020) doi:10.1016/j.scitotenv.2020.138764.

11. Medema, G., Heijnen, L., Elsinga, G., Italiaander, R. & Brouwer, A. Presence of SARS-Coronavirus-2 RNA in Sewage and Correlation with Reported COVID-19 Prevalence in the Early Stage of the Epidemic in The Netherlands. Environ. Sci. Technol. Lett. 7, 511–516 (2020).

12. Crits-Christoph, A. et al. Genome sequencing of sewage detects regionally prevalent SARS-CoV-2 variants. medRxiv 2020.09.13.20193805 (2020) doi:10.1101/2020.09.13.20193805.

13. Kitajima, M. et al. SARS-CoV-2 in wastewater: State of the knowledge and research needs. Science of The Total Environment 739, 139076 (2020).

14. La Rosa, G., Bonadonna, L., Lucentini, L., Kenmoe, S. & Suffredini, E. Coronavirus in water environments: Occurrence, persistence and concentration methods -A scoping review. Water Research 179, 115899 (2020).

15. Lu, D., Huang, Z., Luo, J., Zhang, X. & Sha, S. Primary concentration – The critical step in implementing the wastewater based epidemiology for the COVID-19 pandemic: A mini-review. Science of The Total Environment 747, 141245 (2020).

16. Shortage of RNA extraction kits hampers efforts to ramp up COVID-19 coronavirus testing. Chemical & Engineering News https://cen.acs.org/analytical-chemistry/diagnostics/Shortage-RNA-extraction-kits-hampers/98/web/2020/03.

17. CDC. Coronavirus Disease 2019 (COVID-19). Centers for Disease Control and Prevention https://www.cdc.gov/coronavirus/2019-ncov/cases-updates/wastewater-surveillance/testing-methods.html (2020).

18. Kitajima, M., Sassi, H. P. & Torrey, J. R. Pepper mild mottle virus as a water quality indicator. npj Clean Water 1, 1–9 (2018).

19. Wang, X.-W. et al. Concentration and detection of SARS coronavirus in sewage from Xiao Tang Shan Hospital and the 309th Hospital. Journal of Virological Methods 128, 156–161 (2005).

20. C. Wetter, M. Conti, D. Altschuh, R. Tabillon, and M.H.V. van Regenmortel. Pepper Mild Mottle Virus, a Tobamovirus Infecting Pepper Cultivars in Sicily. Phytopathology (1983).

21. Bivins, A. et al. Persistence of SARS-CoV-2 in Water and Wastewater. Environ. Sci. Technol. Lett. acs.estlett.0c00730 (2020) doi:10.1021/acs.estlett.0c00730.

22. Ahmed, W. et al. Decay of SARS-CoV-2 and surrogate murine hepatitis virus RNA in untreated wastewater to inform application in wastewater-based epidemiology. Environmental Research 191, 110092 (2020).

23. Phillips, S. J., Dalgarn, D. S. & Young, S. K. Recombinant DNA in Wastewater: pBR322 Degradation Kinetics. Research Journal of the Water Pollution Control Federation 61, 1588–1595 (1989).

24. Michael-Kordatou, I., Karaolia, P. & Fatta-Kassinos, D. Sewage analysis as a tool for the COVID-19 pandemic response and management: the urgent need for optimised protocols for SARS-CoV-2 detection and quantification. J Environ Chem Eng 8, 104306 (2020).

25. Schrader, C., Schielke, A., Ellerbroek, L. & Johne, R. PCR inhibitors – occurrence, properties and removal. Journal of Applied Microbiology 113, 1014–1026 (2012).

26. Cao, Y., Griffith, J. F., Dorevitch, S. & Weisberg, S. B. Effectiveness of qPCR permutations, internal controls and dilution as means for minimizing the impact of inhibition while measuring Enterococcus in environmental waters. Journal of Applied Microbiology 113, 66–75 (2012).

27. Pecson, B. M. et al. Reproducibility and sensitivity of 36 methods to quantify the SARS-CoV-2 genetic signal in raw wastewater: findings from an interlaboratory methods evaluation in the U.S. medRxiv 2020.11.02.20221622 (2020) doi:10.1101/2020.11.02.20221622.

28. Whitney, O. Direct wastewater RNA capture and purification via the “Sewage, Salt, Silica and SARS-CoV-2 (4S)” method. (2020) doi:10.17504/protocols.io.biwekfbe.

29. Whitney, O. Direct wastewater RNA extraction via the “Milk of Silica (MoS)” method -A companion method to “Sewage, Salt, Silica and SARS-CoV-2 (4S)” (2020) doi:10.17504/protocols.io.biwfkfbn.

30. Graham, T. G. W. et al. Inexpensive, versatile and open-source methods for SARS-CoV-2 detection. http://medrxiv.org/lookup/doi/10.1101/2020.09.16.20193466 (2020) doi:10.1101/2020.09.16.20193466.

31. Haramoto, E. et al. Occurrence of Pepper Mild Mottle Virus in Drinking Water Sources in Japan. Appl. Environ. Microbiol. 79, 7413–7418 (2013).

32. Decaro, N. et al. Detection of bovine coronavirus using a TaqMan-based real-time RT-PCR assay. Journal of Virological Methods 151, 167–171 (2008).

33. Bustin, S. A. et al. The MIQE Guidelines: Minimum Information for Publication of Quantitative Real-Time PCR Experiments. Clin Chem 55, 611–622 (2009).

34. Wu, F. et al. SARS-CoV-2 Titers in Wastewater Are Higher than Expected from Clinically Confirmed Cases. 5, 9 (2020).

35. Haramoto, E., Malla, B., Thakali, O. & Kitajima, M. First environmental surveillance for the presence of SARS-CoV-2 RNA in wastewater and river water in Japan. Sci Total Environ 737, 140405 (2020).

36. Alameda County COVID-19 Daily Cumulative Cases by City, Place, and Zip Code. https://data.acgov.org/datasets/5d6bf4760af64db48b6d053e7569a47b_3?page=10.

