## Supplementary Information for "Sewage, Salt, Silica and SARS-CoV-2 (4S): An economical kit-free method for direct capture of SARS-CoV-2 RNA from wastewater"

**Supporting Information**

**Supplementary figures**

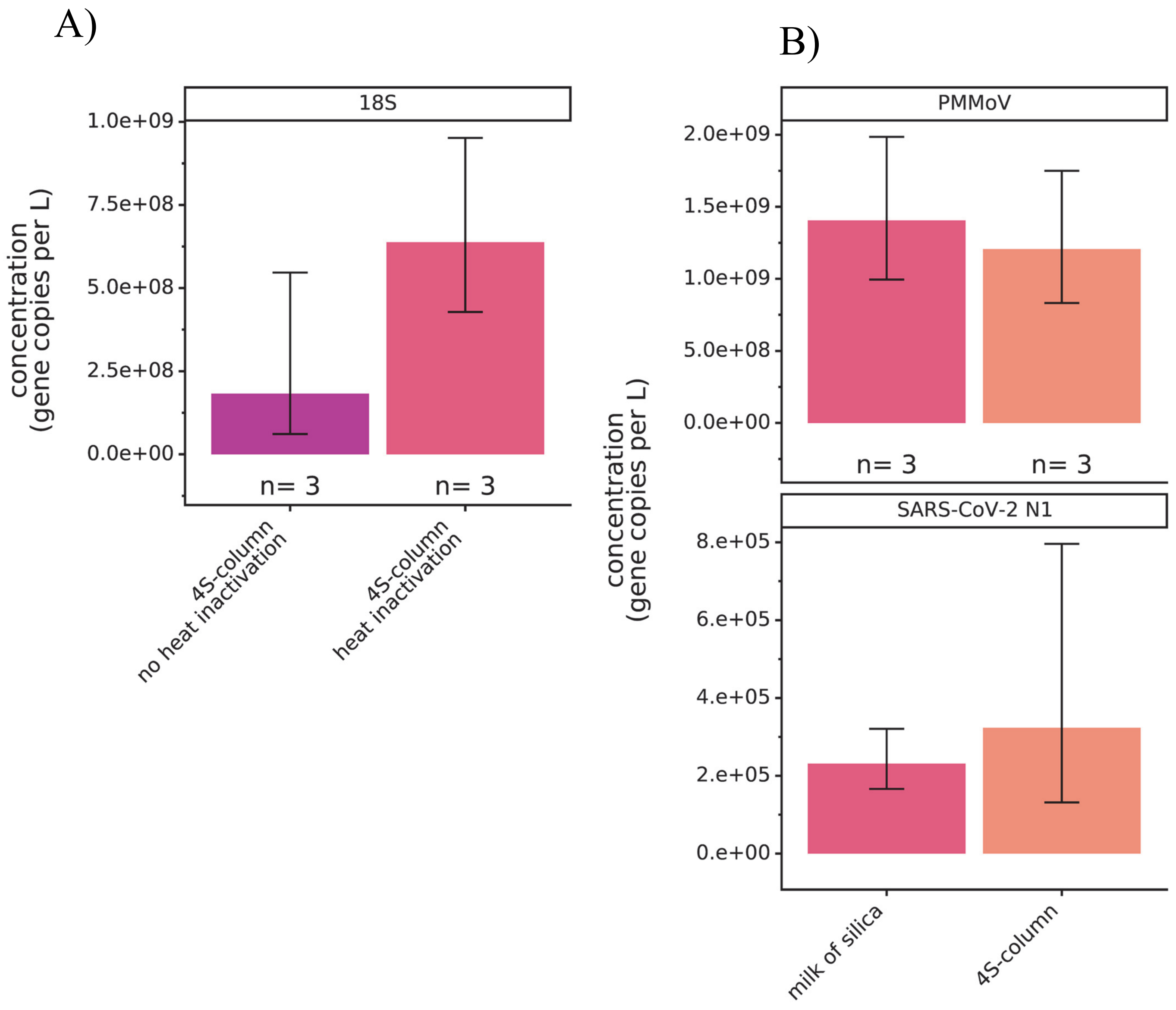


**Figure 1.** A) 18S rRNA assay signal of wastewater RNA purified via the 4S method, with and without heat inactivation. B) SARS-CoV-2 N1 assay and PMMoV assay signal of wastewater RNA purified via the 4S method using “Milk-of-Silica” silicon dioxide particulate (milk of silica) or silica-containing spin columns (4S-column). “n” represents the number of wastewater RNA extraction replicates per condition.

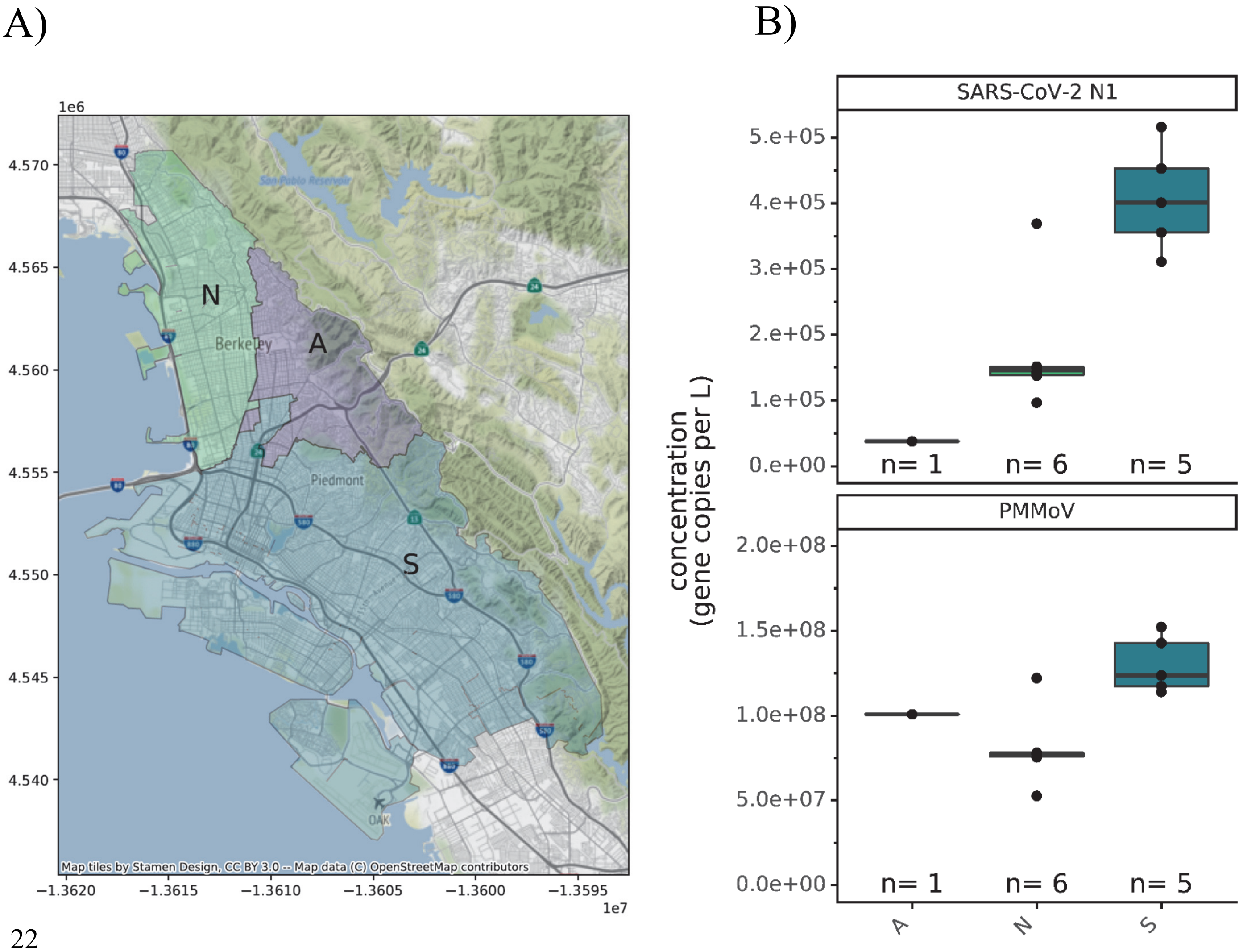


**Figure S2.** A) Sewersheds served by the East Bay Municipal Utility District N, A, and S interceptors. B) SARS-CoV-2 N1 assay and PMMoV assay signal of wastewater RNAs extracted from the A, N and S interceptors. “n” represents the number of wastewater RNA extraction replicates per assay.

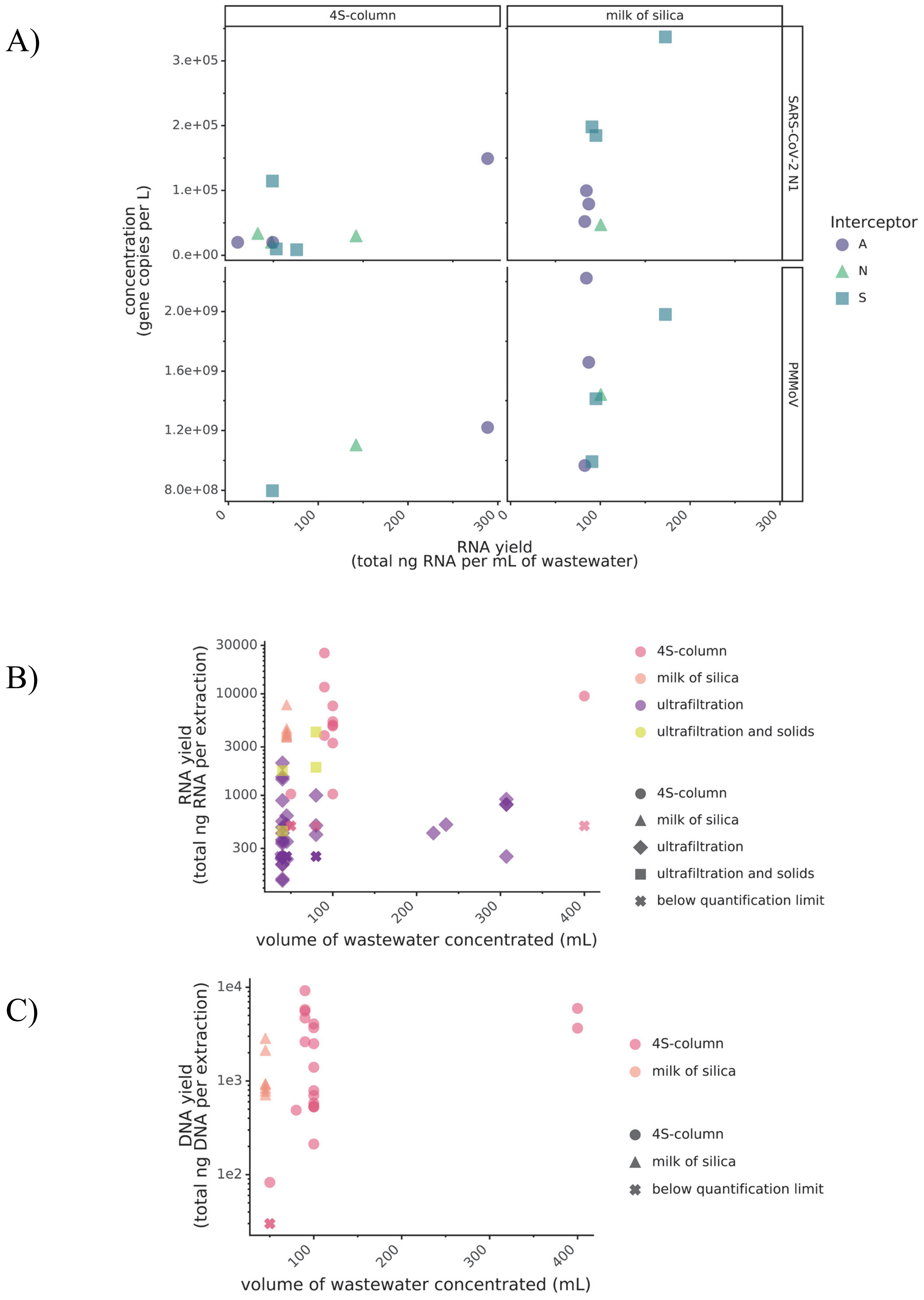
A)
4S-column
milk of silica
SARS-CoV-2 N1
PMMoV
3.e+05
2.e+05
1.e+05
0.e+00
2.0e+09
1.6e+09
1.2e+09
8.0e+08
concentration
(gene copies per L)
0
100
200
300
RNA yield
(total ng RNA per mL of wastewater)
Interceptor
A
N
S
B)
30000
10000
3000
1000
300
RNA yield
(total ng RNA per extraction)
100
200
300
400
volume of wastewater concentrated (mL)
4S-column
milk of silica
ultrafiltration
ultrafiltration and solids
4S-column
milk of silica
ultrafiltration
ultrafiltration and solids
below quantification limit
C)
1e4
1e3
1e2
DNA yield
(total ng DNA per extraction)
100
200
300
400
volume of wastewater concentrated (mL)
4S-column
milk of silica
4S-column
milk of silica
below quantification limit

**Figure S3.** A) Relation of extracted RNA concentration to SARS-CoV-2 N1 and PMMoV assay signal reported as concentration in gene copies per liter, using the 4S-column or Milk-of silica method. X-axis represents RNA yield, defined as ng of RNA extracted per mL of wastewater input.
B) Relationship between wastewater input and RNA yield using 4 different methods, defined as total ng of RNA per extraction. Column (pink) represents the 4S method using a silica column, Milk of Silica (salmon) represents the 4S method using a silicon dioxide binding matrix, ultrafiltration (purple) represents Amicon ultrafilter concentration and ultrafiltration and solids represents Amicon ultrafilter concentration with subsequent RNA extraction of isolated solids.
C) Relationship between wastewater input volume and DNA yield, defined as total ng of DNA per extraction. Column (pink) represents the 4S method using a silica column and Milk of Silica (salmon) represents the 4S method using a silicon dioxide binding matrix.

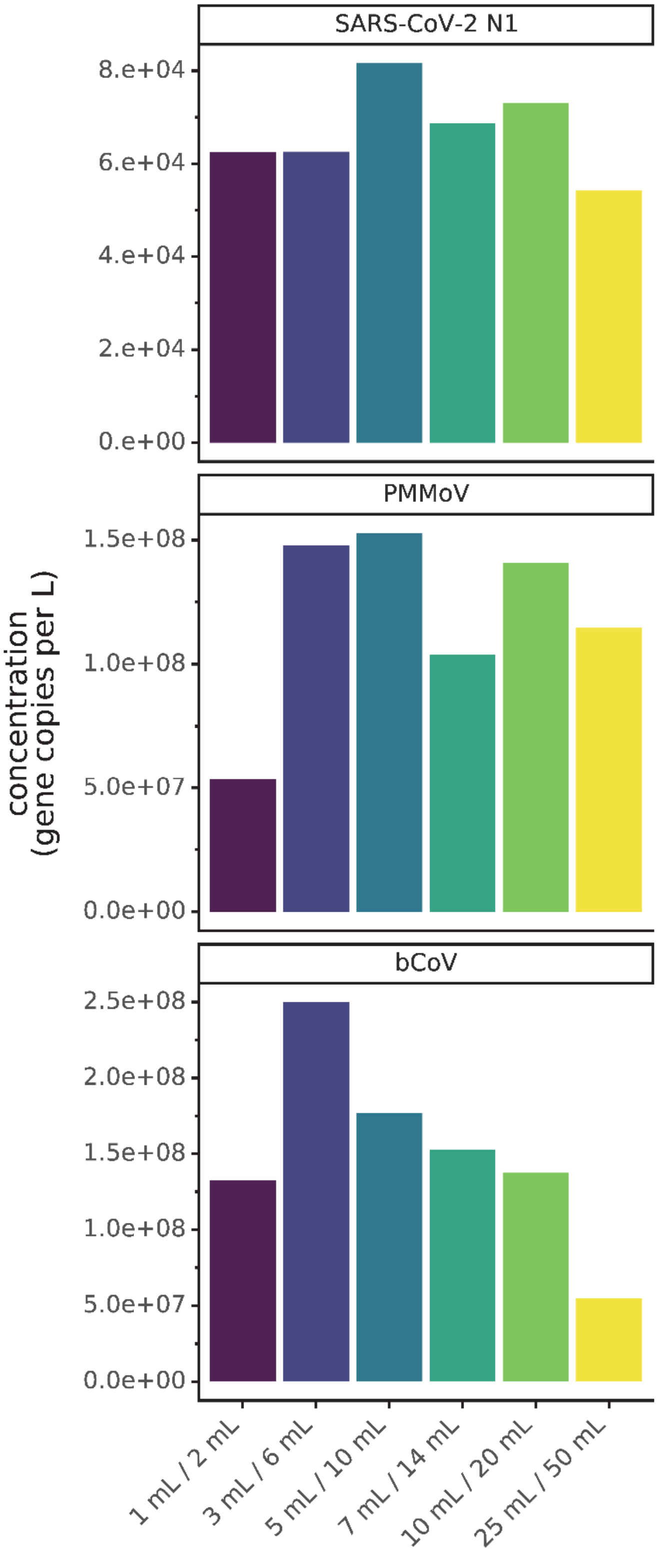


**Figure S4.** Impact of wash buffer volume use on SARS-CoV-2 N1, PMMoV and bCoV assay detection, reported in gene copies per liter. Volumes of wash buffer #1 and wash buffer #2 indicated on x-axis, n=1 extraction replicate per wash buffer volume used.

27

### Supplementary Methods

#### RNA extraction via the 4S "Milk-of-Silica" met

#### Nucleic acid quantification

For all extraction methods, RNA and DNA yields were quantified using the Qubit 4 fluorometer high sensitivity RNA, broad range RNA and broad range DNA quantification assays, following manufacturer instructions. For part of the experiment shown in Supplementary Figure S3A, the solids pellets from slow centrifugation were resuspended in the ultrafiltrate concentrates and used as input for RNA extraction prior to nucleic acid quantification.

#### Target RNA detection via RT-qPCR

Quantification of target RNA species was performed using reverse transcription quantitative polymerase chain reaction (RT-qPCR) in technical triplicate with a QuantStudio 3 Real-Time PCR System (ThermoFisher Scientific). Each 20-μL reaction included extracted RNA (5 μL) and TaqMan Fast Virus 1-Step Master Mix (ThermoFisher Scientific), unless otherwise stated, as well as primers, probes, and RNase/DNase-free water (Table S1). This study employed four primer and probe sets (Table S3; Integrated DNA Technologies) targeting the SARS-CoV-2 nucleocapsid N gene (N1; CDC RUO kit), bovine coronavirus transmembrane protein gene (bCoV; custom DNA oligos ), Pepper mild mottle virus coat protein gene (PMMoV; custom DNA oligos) and human 18S ribosomal rRNA (18S; custom DNA oligos).[1,2] Assay thermocycling conditions are detailed in Table S2, and primer sequence information is in Table S3. Each plate included triplicate no template controls (NTCs) and RNA standards in 10-fold serial dilutions from different manufacturers: for the N1 Assay, synthetic SARS-CoV-2 RNA was used (Control 2- 102024, Twist Bioscience, San Francisco, CA); for bCoV and PMMoV, custom Ultramer RNA Oligonucleotides were used (Integrated DNA Technologies); and for 18S, RNA was in-vitro transcribed from an 18S target amplicon geneBlock (Integrated DNA Technologies) using the HiScribe T7 Quick High Yield RNA Synthesis Kit (New England Biolabs). Throughout all experiments, two no template controls amplified, but both were at least 8 Cq higher than that of the lowest point on the standard curve, and standard curve efficiencies ranged from 75% to 105%. Quality assessment data (including efficiencies and $R^2$ values by plate) are detailed in the supporting information (Table S4). Inhibition testing was completed once for each wash buffer condition for the N1, bCoV, and PMMoV assays following the spike and dilute method (Figure 4).[3] A MIQE guideline checklist for this manuscript can be found in table S6.[4]

**Data analysis**

qPCR technical triplicate Cq values were determined through automatic thresholding on QuantStudio 3 Design and Analysis Software (v1.5.1). These values were imported into a custom data analysis pipeline in python (v3.6.9) which utilized key modules for manipulation (Pandas v1.1.2), calculations (NumPy v1.18.5 and SciPy v1.4.1), and visualizations (plotnine v0.6.0). First, for RT-qPCR technical triplicates, outlier identification and disposition were performed using the two-sided Grubbs test (with alpha = 0.02505) in the outlier-utils module (v0.03; Table S5). After outlier removal, the geometric mean of technical replicates was determined for samples and standards with two or more technical replicates. To adjust for fecal concentration, a normalized value was calculated by dividing the quantity (gc/L) of N1 by the associated PMMoV quantity (gc/L) for comparisons with population COVID-19 case data.[5,6] Linear regression was performed (scikit-learn v0.22.2) using plate-specific standard curves, unless otherwise stated, to determine associated quantity in gene copies per reaction in samples. Plates 35 and 38 did not have associated standard curves, thus the standard curve from plate 32 was used (all bCoV plates). Samples were considered below the limit of quantification if the Cq was higher than the Cq of the lowest quantity on the standard curve of the plate, or if the Cq was greater than 40. One hundred percent of replicates used in standard curves for 18S and PMMoV were detected. However, for N1 and bCoV, the limit of detection was set to be 20 and 1000 gene copies per reaction, respectively; the resulting percentage of detectable replicates was greater than 90% (Table S5). Thirteen samples were deemed below the limit of quantification or below the limit of detection; the limit of detection was used for these samples for plotting and data analysis. The quantity values were converted to gene copies per liter of wastewater for test group comparisons.

The geometric mean and geometric standard deviation of biological replicates was determined, and all error bars in this manuscript represent the geometric standard deviation range of biological replicates with number of replicates (n) shown on each plot. Normalized N1 to PMMoV values as well as gene copy per L data for all targets were not normally distributed, thus the statistical method used to assess results significance in Figure 2 was the Kruskal Wallis nonparametric test.

To compare wastewater results with case data, publicly available new cases per day data from Alameda county were used.[7] To estimate new cases per day in each sub-sewershed, shape files of the A, N, and S sub-sewersheds (Figure S1) and of zip codes in California were overlayed in GeoPandas (v0.8.1). The fraction of overlap between each zip code and each interceptor was used to calculate the area-averaged new cases per day in each sub-sewershed.

Associated data used to generate each figure is included in the supplemental files.

### Tables

Table S1: Reaction conditions for each assay

| Reaction Component | N1 Reaction concentration (µM) | PMMoV Reaction concentration (µM) | bCoV Reaction concentration (µM) | 18S Reaction concentration (µM) |
|---|---|---|---|---|
| TaqMan Fast Virus 1-Step Master Mix* | 1x | 1x | 1x | 1x |
| Primer F | 0.5 | 0.4 | 0.9 | 0.5 |
| Primer R | 0.5 | 0.4 | 0.9 | 0.5 |
| Probe | 0.12 | 0.2 | 0.25 | 0.13 |

*Plates 12, 16, and 17 used TaqPath 1-Step RT-qPCR Master Mix at the same concentration

Table S2: Thermocycling parameters for all assays

| Thermocycling conditions | | |
|---|---|---|
| **Reaction Cycling Step** | **Temperature (°C)** | **Time (minutes: seconds)** |
| **UNG incubation** | 25 | 2:00 |
| **RT step** | 50 | 15:00 |
| **Polymerase activation** | 95 | 2:00 |
| **45 cycles** | 95 | 0:03 |
| | 55 | 0:30 |

Table S3: qPCR assay information for the SARS-CoV-2 nucleocapsid N gene (N1), the bovine coronavirus transmembrane protein gene (bCoV), the pepper mild mottle virus coat protein gene (PMMoV) and human 18S ribosomal rRNA (18S)

| **Gene target** | **Type of Sequence (length; accession)** | **Sequence (5' -> 3')** |
| --- | --- | --- |
| N1 | Forward primer | GACCCCAAAATCAGCGAAAT |
| | Reverse primer | TCTGGTTACTGCCAGTTGAATCTG |
| | Probe | FAM-ACCCCGCATTACGTTTGGTGGACC- MGB-NFQ |
| | Amplicon (72 bp; MN908947.3) | GACCCCAAAATCAGCGAAATGCACCCCGCATTACGTTTGGTGGACCCTCAGATTCAACTGGCAGTAACCAGA |
| PMMoV | Forward primer | GAGTGGTTTGACCTTAACGTTTGA |
| | Reverse primer | TTGTCGGTTGCAATGCAAGT |
| | Probe | FAM-CCTACCGAAGCAAATG-MGB-NFQ |
| | Amplicon (68 bp; AB716964) | GAGTGGTTTGACCTTAACGTTTGAGCGGCCTACCGAAGCAAATGTCGCACTTGCATTGCAACCGACAA |
| bCoV | Forward primer | CTGGAAGTTGGTGGAGTT |
| | Reverse primer | ATTATCGGCCTAACATACATC |
| | Probe | FAM-CCTTCATATCTATACACATCAAGTTGTT- MGB-NFQ |
| | Amplicon (85 bp; AF39154) | CTGGAAGTTGGTGGAGTTTCAACCCAGAAACAAACAACTTGATGTGTATAGATATGAAGGGAAGGATGTATGTTAGGCCGATAAT |
| 18S | Forward primer | GGTTCCTTTGGTCGCTCGCT |
| | Reverse primer | GGGCTGACCGGGTTGGTTTT |
| | Probe | /56-FAM/AG AGC TAA T/ZEN/A CAT GCC GAC GGG C/3IABkFQ/ |
| | Amplicon (138bp; 6G18_2) | GGTTCCTTTGGTCGCTCGCTCCTCTCCTACTTGGATAACTGTGGTAATTCTAGAGCTAATACATGCCGACGGGCGCTG ACCCCCTTCGCGGGGGGGATGCGTGCATTTATCAGATCAAAACCAACCCGGTCAGCCC |

Table S4: RT-qPCR validation information

| plate id | Target | linear dynamic range (orders of magnitude) | lowest quantity on the standard curve (Cq) | lowest quantity on the standard curve (gene copies per L) | lowest quantity on the standard curve (geometric standard deviation) | slope | y-intercept | $R^2$ | PCR efficiency | Minimum Cq of NTC triplicates | intraassay variation (arithmetic mean of coefficient of variation of quantities on each plate) |
|---|---|---|---|---|---|---|---|---|---|---|---|
| **12** | N1 | 5 | 34.81 | 10 | 1.013 | -3.21 | 38.57 | 0.9917 | 1.05 | negative | 2.602 |
| **16** | N1 | 5 | 36.04 | 20 | 1.004 | -3.59 | 40.99 | 0.9977 | 0.898 | negative | 4.617 |
| **17** | PMMoV | 6 | 35 | 1000 | 1.004 | -3.61 | 45.62 | 0.9987 | 0.892 | negative | 16.198 |
| **20** | N1 | 6 | 34.93 | 10 | 1.013 | -3.49 | 38.38 | 0.9983 | 0.935 | negative | 3.515 |
| **21** | N1 | 5 | 35.53 | 10 | 1.007 | -3.5 | 39.2 | 0.9991 | 0.932 | negative | 1.642 |
| **22** | PMMoV | 6 | 35.01 | 1000 | 1.007 | -3.81 | 47.38 | 0.9923 | 0.829 | negative | 5.037 |
| **23** | PMMoV | 6 | 35.59 | 1000 | 1.013 | -4.13 | 49.84 | 0.9494 | 0.745 | negative | 2.543 |
| **27** | N1 | 5 | 34.46 | 20 | 1.018 | -3.56 | 38.81 | 0.9969 | 0.909 | negative | 2.546 |
| **28** | PMMoV | 4 | 29.52 | 10000 | 1.006 | -3.44 | 43.12 | 0.9984 | 0.955 | 39.0265444 | 8.882 |
| **32** | bCoV | 6 | 37.89 | 100 | 1.014 | -3.79 | 46.07 | 0.9956 | 0.837 | negative | 6.506 |
| **34** | N1 | 6 | 36.84 | 10 | 1.002 | -3.32 | 40.21 | 0.9992 | 1.001 | negative | 2.171 |
| **35** | bCoV | 6 | 37.89 | 100 | 1.014 | -3.79 | 46.07 | 0.9956 | 0.837 | negative | 1.532 |
| **36** | PMMoV | 6 | 36.06 | 1000 | 1.006 | -4.29 | 50.4 | 0.9705 | 0.71 | negative | 1.274 |
| **37** | N1 | 5 | 35.19 | 20 | 1.003 | -3.44 | 39.74 | 0.9997 | 0.953 | negative | 0.152 |
| **38** | bCoV | 6 | 37.89 | 100 | 1.014 | -3.79 | 46.07 | 0.9956 | 0.837 | negative | 0.659 |
| **39** | PMMoV | 6 | 34.46 | 1000 | 1.004 | -3.86 | 47.03 | 0.9842 | 0.816 | negative | 0.485 |
| **59** | 18S | 6 | 29.36 | 1000 | 1.009 | -3.42 | 39.61 | 0.9999 | 0.962 | 37.7260635 | 9.094 |

Table S5: Evidence for LoD/LoQ

| assay | standard curve Quantity (gene copies per reaction) | fraction of replicates positive | number of replicates that passed Grubbs test | number of replicates that failed Grubbs test | total number of replicates |
|---|---|---|---|---|---|
| **N1** | 10 | 0.75 | 12 | 9 | 21 |
| **N1** | 20 | 0.923 | 13 | 5 | 18 |
| **N1** | 100 | 1 | 16 | 5 | 21 |
| **N1** | 1000 | 1 | 18 | 3 | 21 |
| **N1** | 10000 | 1 | 17 | 4 | 21 |
| **N1** | 100000 | 1 | 17 | 4 | 21 |
| **PMMoV** | 1000 | 1 | 12 | 3 | 15 |
| **PMMoV** | 10000 | 1 | 13 | 5 | 18 |
| **PMMoV** | 100000 | 1 | 16 | 2 | 18 |
| **PMMoV** | 1000000 | 1 | 14 | 4 | 18 |
| **PMMoV** | 10000000 | 1 | 14 | 4 | 18 |
| **PMMoV** | 100000000 | 1 | 11 | 4 | 15 |

| | | | | | |
|---|---|---|---|---|---|
| **bCoV** | 100 | 0.667 | 3 | 0 | 3 |
| **bCoV** | 1000 | 1 | 2 | 1 | 3 |
| **bCoV** | 10000 | 1 | 2 | 1 | 3 |
| **bCoV** | 100000 | 1 | 3 | 0 | 3 |
| **bCoV** | 1000000 | 1 | 2 | 1 | 3 |
| **bCoV** | 10000000 | 1 | 3 | 0 | 3 |
| **18S** | 1000 | 1 | 2 | 1 | 3 |
| **18S** | 10000 | 1 | 2 | 1 | 3 |
| **18S** | 100000 | 1 | 3 | 0 | 3 |
| **18S** | 1000000 | 1 | 2 | 1 | 3 |
| **18S** | 10000000 | 1 | 2 | 1 | 3 |
| **18S** | 100000000 | 1 | 3 | 0 | 3 |

Table S6: Reagent cost of the 4S method using a silica column or silicon dioxide particulate

| **Material** | **4S-Column cost** | **4S “Milk-of-Silica” cost** |
|---|---|---|
| 50 mL tube | $0.36 | $0.18 |
| NaCl | $0.48 | $0.48 |
| Tris | $0.02 | $0.02 |
| EDTA | $0.02 | $0.02 |
| Bovilis Coronavirus Calf Vaccine | $0.04 | $0.04 |
| 50 mL syringe | $0.78 | $0.78 |
| 5uM 47mm PVDF membrane filter | $1.56 | $1.56 |
| Silicon Dioxide | N.A. | $0.62 |
| Zymo IIIP columns with reservoir | $8.41 | N.A. |
| Ethanol (70%) | $0.56 | $0.57 |
| Ethanol (96%) | $0.13 | $0.13 |
| 1.5 mL tube | $0.02 | $0.02 |
| 1.5 mL Lo-bind tube | $0.13 | $0.13 |

| | | |
|---|---|---|
| PCR water | NA | $1.82 |
| Isopropanol | NA | $1.18 |
| Sodium Acetate | NA | $0.06 |
| Elution Buffer (ZymoPURE) | $0.31 | $0.31 |
| Total Cost | $12.83 | $7.93 |

Table S7: MIQE guideline essential information checklist

| Requirement for submission | Requirement or location of requirement |
|---|---|
| **Experimental Design** | |
| Definition of experimental and control groups | Experimental groups defined as wastewaters purified via different extraction methods, with or without the addition of preservative salts. |
| Number within each group | Three wastewater RNA extraction replicates of an influent sample within each experimental group |
| **Sample** | |
| Description | Wastewater influent sample |
| Microdissection or macrodissection | Not applicable |
| Processing procedure | 4S or ultrafiltration-based RNA extraction |
| If frozen, how quickly | Not applicable |
| If fixed, with what and how quickly | Not applicable |
| Sample storage conditions and duration | Samples stored for 1-2 days at 4°C or <2 weeks at -80°C |
| **Nucleic Acid Extraction** | |
| Procedure/instrumentation | "4S" purification using silica columns or silica particulate, Ultrafiltration as previously described, with subsequent AllPrep kit RNA extraction |
| Name of kit and details of modifications | Amicon 100kDA ultrafilter, AllPrep RNA/DNA extraction kit |
| Details of DNase or RNase treatment | Not applicable |
| Contamination assessment (DNA or RNA) | Not applicable |
| Nucleic acid quantification | Fluorescent dyes reactive to RNA and DNA |
| Instrument and method | Qubit4 fluorometer, high-sensitivity and broad-range RNA assay, broad-range DNA assay |
| RNA integrity: method/instrument | Not applicable |
| RIN/RQI or $C_q$ of 3′ and 5′ transcripts | Not applicable |
| Inhibition testing | Spike and dilute method; RT-qPCR methods section |
| **Reverse Transcription** | |
| Complete reaction conditions | Table S1 |
| Amount of RNA and reaction volume | 5µL of extracted RNA and 20 µL reaction volume |
| Priming oligonucleotide and concentration | 0.5uM- 0.9uM, detailed in table S1 |

| | |
|---|---|
| Reverse transcriptase concentration | Included in TaqMan Fast Virus 1-Step Master Mix and TaqPath 1-Step RT-qPCR Master Mix (ThermoFisher Scientific) |
| Temperature and time | Table S2 |
| **qPCR target information** | |
| Gene symbol | N – SARS-CoV-2 Nucleocapsid, RNA18S1 – RNA, 18S ribosomal 1, M – Membrane protein, bovine coronavirus, CP – PMMoV Coat protein |
| Sequence accession number | Table S3 |
| Amplicon length | Table S3 |
| In silico specificity screen | NCBI Primer-BLAST |
| Location of each primer by exon or intron | Not applicable |
| What splice variants are targeted | Not applicable |
| **qPCR oligonucleotides** | |
| Primer Sequences | Table S3 |
| Location and identity of any modifications | Not applicable |
| **qPCR protocol** | |
| Complete reaction conditions | Table S1 |
| Reaction volume and amount of cDNA | 20 µL, Not Applicable (1-Step RT-qPCR) |
| Primer, probe, $Mg^{2+}$, and dNTP concentrations | Included in TaqMan Fast Virus 1-Step Master Mix and TaqPath 1-Step RT-qPCR Master Mix (ThermoFisher Scientific) |
| Polymerase identity and concentration | Included in TaqMan Fast Virus 1-Step Master Mix and TaqPath 1-Step RT-qPCR Master Mix (ThermoFisher Scientific) |
| Kit identity and manufacturer | RT-qPCR methods section |
| Additives | Not applicable |
| Complete thermocycling parameters | Table S2 |
| Manufacturer of qPCR instrument | QuantStudio 3 Real-Time PCR system (ThermoFisher Scientific) |
| **qPCR validation** | |
| Specificity | No detected amplification of assays within negative RT-qPCR controls |
| For SYBR Green I, $C_q$ of the NTC | Not applicable |
| Calibration curves with slope and y intercept | Table S4 |
| PCR efficiency | Table S4 |
| $R^2$ | Table S4 |
| Linear dynamic range | Table S4 |
| $C_q$ variation at LoD | Table S4 |
| Evidence for LoD | Table S5 |
| If multiplex, efficiency and LoD of each assay | Not applicable |
| **Data Analysis** | |
| qPCR analysis program | Custom Python pipeline; Data analysis methods section |

| | |
|---|---|
| Method of $C_q$ determination | Automatic thresholding through Design and Analysis Software v1.5.1 |
| Outlier identification and disposition | Grubbs test; Data analysis methods section |
| Results for NTCs | Table S4 |
| Justification of number and choice of reference genes for normalization method | Not applicable |
| Description of normalization method | Not applicable |
| Number and stage of technical replicates | RT-qPCR technical triplicates |
| Repeatability | Table S4 |
| Statistical methods for results significance | Data analysis methods section |
| Software | Firmware v1.3.3, QuantStudio 3;<br>Design and Analysis Software v1.5.1; QuantStudio 3 |

**<u>References</u>**


1. Haramoto, E. *et al.* Occurrence of Pepper Mild Mottle Virus in Drinking Water Sources in Japan. *Appl. Environ. Microbiol.* **79**, 7413–7418 (2013).
2. Decaro, N. *et al.* Detection of bovine coronavirus using a TaqMan-based real-time RT-PCR assay. *Journal of Virological Methods* **151**, 167–171 (2008).
3. Cao, Y., Griffith, J. F., Dorevitch, S. & Weisberg, S. B. Effectiveness of qPCR permutations, internal controls and dilution as means for minimizing the impact of inhibition while measuring Enterococcus in environmental waters. *Journal of Applied Microbiology* **113**, 66–75 (2012).
4. Bustin, S. A. *et al.* The MIQE Guidelines: Minimum Information for Publication of Quantitative Real-Time PCR Experiments. *Clin Chem* **55**, 611–622 (2009).
5. Wu, F. *et al.* SARS-CoV-2 Titers in Wastewater Are Higher than Expected from Clinically Confirmed Cases. **5**, 9 (2020).
6. Haramoto, E., Malla, B., Thakali, O. & Kitajima, M. First environmental surveillance for the presence of SARS-CoV-2 RNA in wastewater and river water in Japan. *Sci Total Environ* **737**, 140405 (2020).

7. Alameda County COVID-19 Daily Cumulative Cases by City, Place, and Zip Code. https://data.acgov.org/datasets/5d6bf4760af64db48b6d053e7569a47b_3?page=10.
